# Genomic, antigenic and transmission dynamics of influenza A(H3N2) subclade K in Canada, early 2025/26 season

**DOI:** 10.64898/2026.02.10.26345998

**Authors:** George S. Long, Thomas W.A. Braukmann, Nicholas Waglechner, Patryk Aftanas, Alex Marchand-Austin, Julianne V. Kus, Shawn T. Clark, Kevin Katz, Maan Hasso, Finlay Maguire, Samir N. Patel, Samira Mubareka, Venkata R. Duvvuri

**Affiliations:** Public Health Ontario, Toronto, ON, Canada; Department of Laboratory Medicine and Pathobiology, Faculty of Medicine, University of Toronto, Toronto, ON, Canada; Sunnybrook Research Institute, Toronto, ON, Canada; Shared Hospital Laboratory, Toronto, ON, Canada; Faculty of Computer Science, Dalhousie University, Halifax, NS, Canada; Department of Community Health & Epidemiology, Faculty of Medicine, Dalhousie University, Halifax, NS, Canada

**Keywords:** Influenza, Subclade K, Antigenic Diversity, Phylodynamics, Phylogeography

## Abstract

Influenza A(H3N2) subclade K virus was detected in Canada early in the 2025/26 influenza season, bearing an antigenic transition in the hemagglutinin (HA) glycoprotein. Analysis of 396 HA sequences from Canada showed antigenic divergence from 2025/26 influenza vaccine strains, consistent with partial mismatch. Phylodynamic analysis revealed sustained pre-vaccine transmission without clear post-vaccine expansion. Phylogenetic and phylogeographic analyses indicated interprovincial mixing within a highly connected metapopulation, highlighting the value of genomic surveillance for real-time epidemiologic inference and public health decision-making.

---

An antigenic transition in the circulating seasonal influenza A(H3N2) virus hemagglutinin (HA) glycoprotein was detected in the southern hemisphere (SH) during the 2024/25 season. This transition, comprising seven amino acid (AA) substitutions (K2N, S144N, N158D, I160K, T328A, Q173R, and S378N), led to the emergence of influenza A(H3N2) subclade J.2.4.1 (renamed subclade K) against a background of circulating J.2.3 and J.2.4 viruses [1,2]. Influenza A(H3N2) subclade K virus has since been reported in the northern hemisphere (NH), with experimental evidence of antigenic mismatch relative to cell- and egg-propagated vaccine strains, appearing more pronounced for the NH 2025/26 egg-propagated vaccine [1–4]. Canadian sentinel surveillance of the early 2025/26 season reported an adjusted vaccine effectiveness (VE) of 38% [27%, 47%] for influenza A and 37% [20%, 50%] for subclade K specifically [5]. Despite these observations, it remains unclear whether the antigenic divergence associated with subclade K translates into altered transmission dynamics. We therefore conducted sequence-based, antigenic, and phylodynamic analyses to characterize the emergence of subclade K and to evaluate how immunogenic changes relate to patterns of viral transmissibility and geographic spread across Ontario and Canada.

## Phylogenetic and Antigenic Divergence Analyses

Early in the 2025/26 influenza season, subclade K viruses from Canada formed a monophyletic clade, with sequences from provinces for which sequence data were available interspersed throughout a time-scaled maximum-likelihood phylogeny inferred from HA alignments (Figure 1; Supplementary Figure S1). Province-specific phylogenies were re-estimated using identical model settings, yielding a mean evolutionary rate for Canada of 3.12×10^−3^ substitutions/site/year. Provincial estimates varied, with rates of 1.27×10^−3^ in British Columbia, 6.67×10^−4^ in Saskatchewan, 2.87×10^−3^ in Manitoba, 5.88×10^−3^ in Ontario, 3.67×10^−3^ in New Brunswick, and 2.58×10^−3^ in Nova Scotia (Supplementary Materials). These estimates were lower than the global subclade K rate, 7.77 × 10^−3^ [2]. We next quantified antigenic level divergence relative to vaccine strains using the FLU substitution model [6]. FLU distance values (Supplementary Figure S2) were used as input for a principal coordinates analysis, which placed subclade K in a distinct region of HA sequence space separated from previous vaccine strains (Figure 2**A**). To assess whether this separation could lead to significant vaccine mismatch, we estimated epitope-level antigenic distance using pEpitope [7] (Figure 2**B**). Relative to previous seasons vaccine strains (groups I-II) (Figure 2**B**, Supplementary Table S1), subclade K showed elevated antigenic distances (pEpitope = 0.23), exceeding thresholds commonly associated with substantial immune escape (pEpitope ≥ 0.20) [7]. Antigenic divergence was reduced relative to vaccine group III (NH 2025/26 and SH 2025; pEpitope = 0.17) and was lowest relative to vaccine group IV (SH 2026; pEpitope = 0.078). Approximately half of amino acid (AA) substitutions (19/35, 54.28%) in HA antigenic sites of circulating influenza A(H3N2) viruses occurred in or within five AA residues of predicted N-linked glycosylation motifs (Supplementary Table S2 and S3). pEpitope profiles were similar across Canadian provinces, with no marked differences in antigenic-site divergence (Supplementary Figure S3).

**Figure 1.**
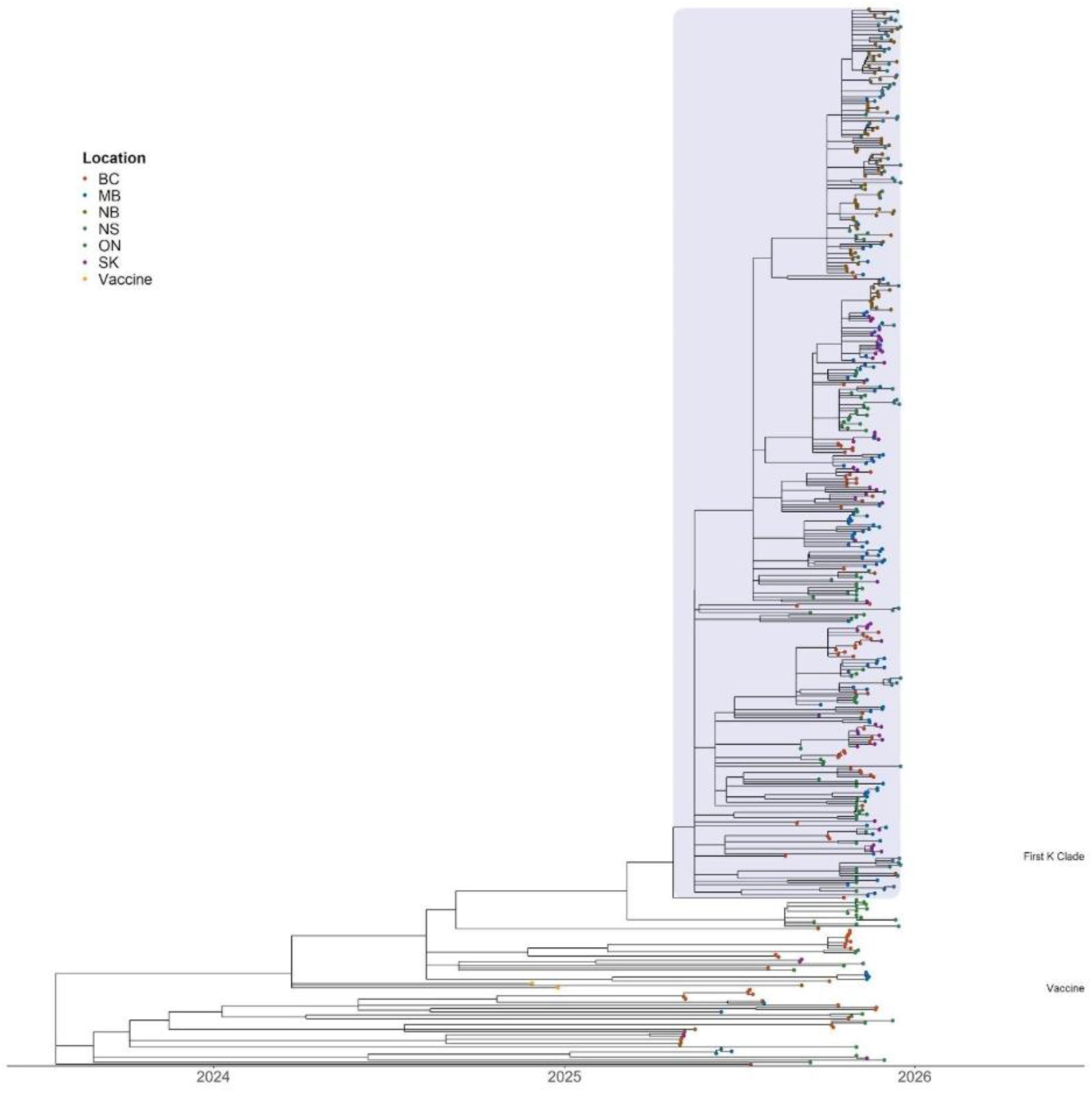
Time-Scaled Phylogeny of Influenza A(H3N2) Subclade K Showing Persistence and Interprovincial Mixing in Canada. Subclade K highlighted in blue. Time scaled maximum-likelihood tree with tips colored by province of sample collection (BC: British Columbia; SK: Saskatchewan; MB: Manitoba; ON: Ontario; NB: New Brunswick and NS: Nova Scotia). Branch lengths represent evolutionary time, and clustering reflects genetic relatedness among viruses. The interspersed distribution of provincial colors throughout the tree indicates frequent interprovincial mixing and limited long-lived, province-specific lineages. Shaded regions highlight periods of sustained circulation of subclade K during the early 2025/26 influenza season (August - December 2025). Indicated on the right side of the tree is the placement the 2026 Southern Hemisphere vaccines strains for egg-based, cell-culture and recombinant-based vaccines, as well as the first observed K-clade sequence in Canada.

**Figure 2.**
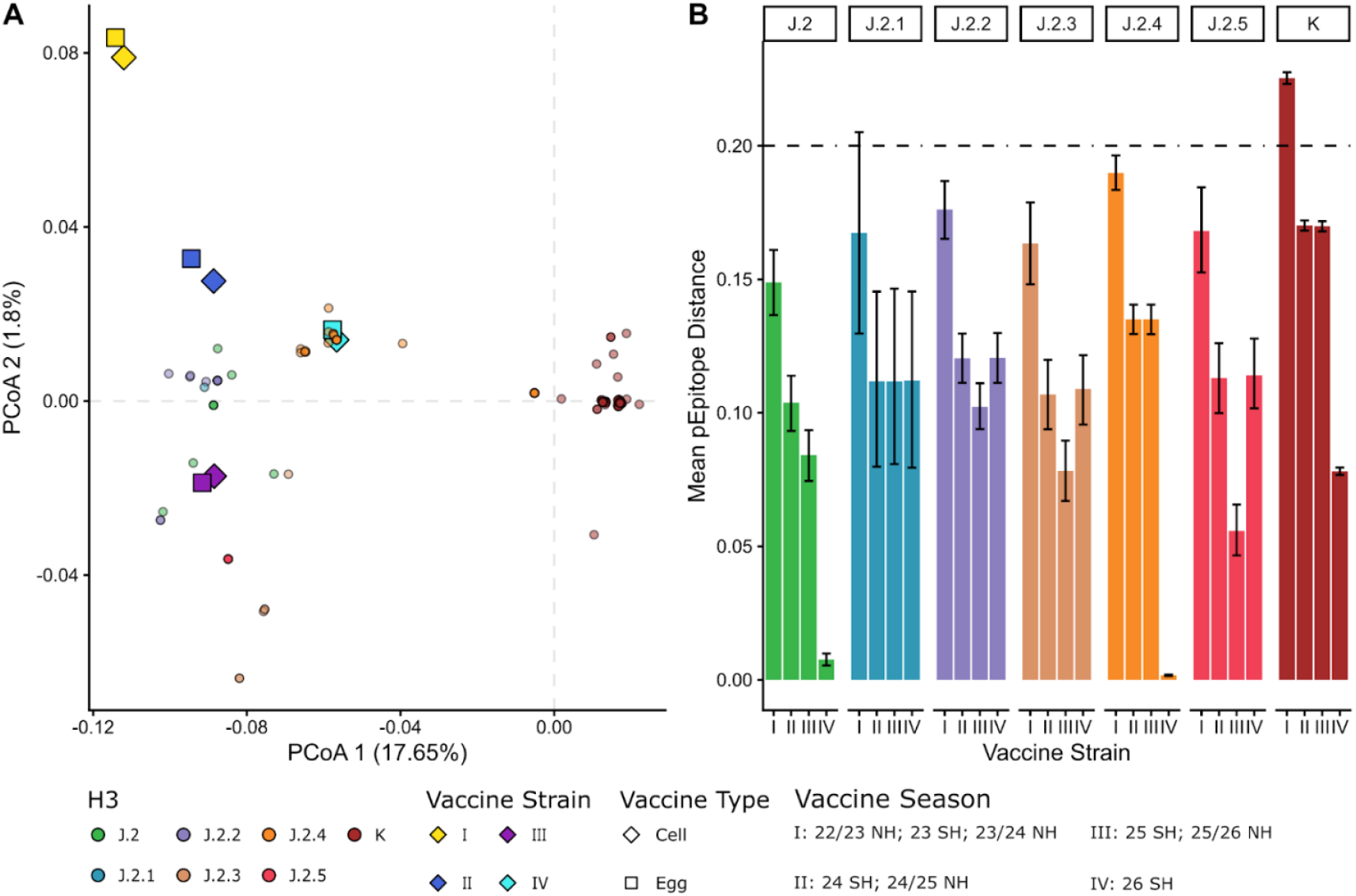
Characterization of circulating influenza A(H3N2) viruses in Canada relative to vaccine strains recommended for Northern and Southern Hemispheres for 2022–2026 seasons. **A)** Principal coordinates analysis (PCoA) of sequence distances based on the FLU substitution matrix. Each point represents a virus or vaccine strain coloured by subclade. Axes show the proportion of variance explained by each principal coordinate. The mean distances can be found in Supplementary Figure S2. **B)** Mean antigenic distance. Bars represent mean pEpitope values with error bars indicating variability across sequences. Black-dashed horizontal line denotes a pEpitope threshold (≥0.20) [7] for increased antigenic divergence; higher values indicate greater vaccine mismatch potential.

## Transmission Dynamics and Spatial Spread of Subclade K viruses

We evaluated whether these phylogenetic and antigenic divergences were associated with altered transmission dynamics and spatial spread using phylodynamic and phylogeographic analyses (Figure 3, Supplementary Methods) with time-stamped HA sequences of subclade K. Using the birth-death skyline phylodynamic model [8], we estimated time-varying reproduction numbers (R_t_) across Canada as well as Ontario, which was selected for its population size, travel connectivity, and genomic surveillance data from two independent organizations, from August to December 2025 (Figure 3**A**, Supplementary Table S4). During the pre-vaccine period, R_t_ estimates clustered near the epidemic threshold for both Canada (R_t_ = 0.9–1.2) and Ontario (R_t_ = 0.8–1.8). Following the staggered introduction of the 2025/26 seasonal influenza vaccine across Canadian provinces between October 13 and October 27, 2025, national R_t_ estimates fluctuated around the epidemic threshold, with alternating periods of lower and higher transmission intensity (R_t_ = 0.78–1.47). In Ontario, where vaccination began on October 27, R_t_ declined thereafter and remained below 1, consistent with reduced transmission. To further contextualize these dynamics, we applied Bayesian phylogeography [9], revealing frequent interprovincial viral movement consistent with a highly connected transmission network (Figure 3**B**).

**Figure 3.**
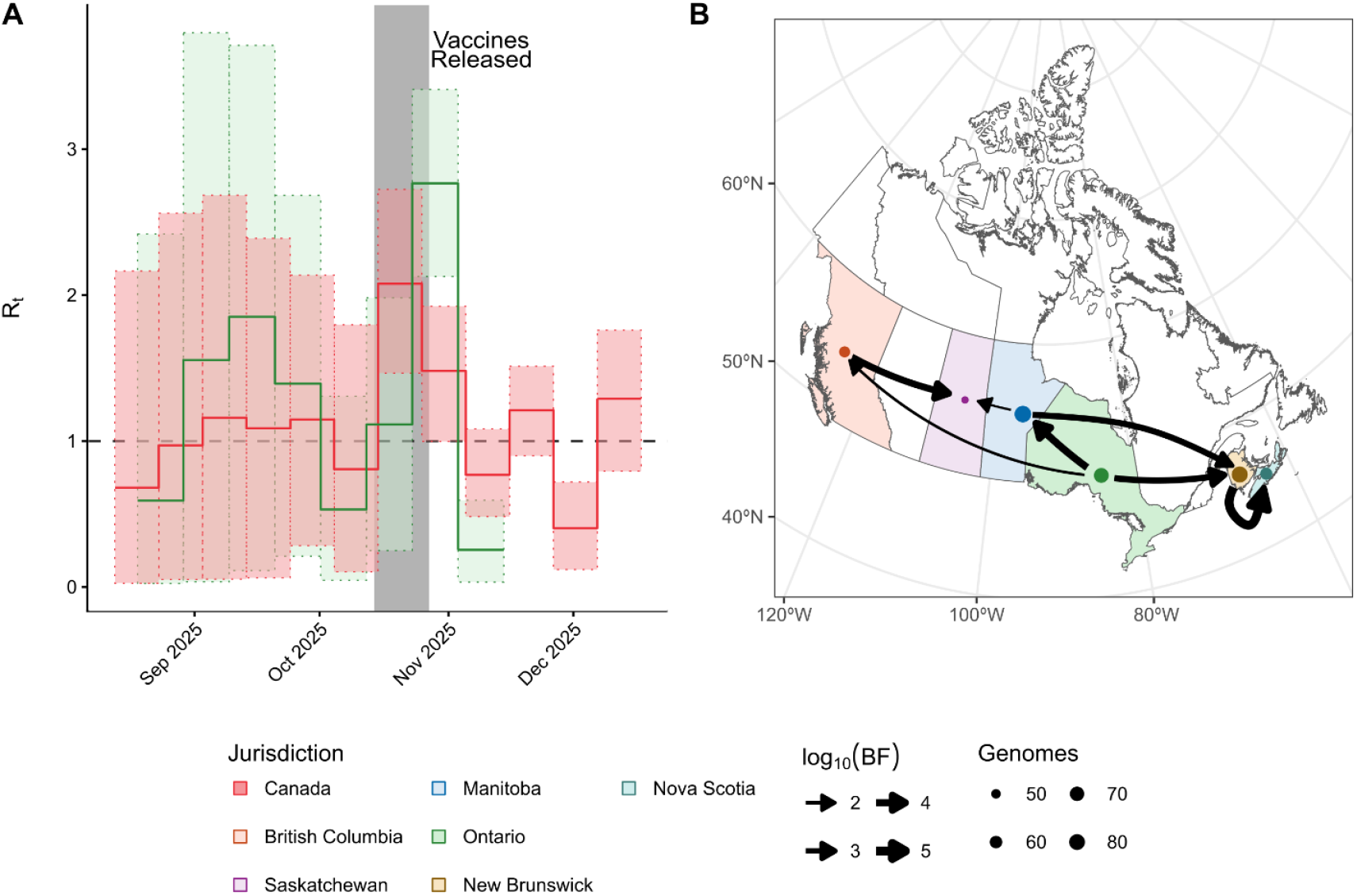
Transmission Potential and Interprovincial Spread Dynamics of Subclade K in Canada. **A)** Time-varying effective reproduction numbers (R_t_) inferred using birth–death skyline phylodynamic models [8] from time-stamped HA sequences. Estimates are shown for Canada (red) and Ontario (green) from August to December 2025. Solid lines indicate median R_t_ estimates and shaded areas represent 95% highest posterior distributions (HPDs). The horizontal dashed line denotes the epidemic threshold (R_t_ = 1), and the vertical grey bar marks the introduction of the 2025/26 seasonal influenza vaccine across Canadian provinces. **B) Genomically inferred interprovincial transmission pathways of subclade K across Canada**, estimated using a Bayesian discrete phylogeographic model [9]. Arrows indicate the direction of inferred viral movement between provinces, with arrow thickness proportional to log_10_ Bayes factor support. Only transmission pathways with moderate (log^10^(BF) > 1) to strong statistical support are shown. Interpretations of the log (BF): 0 to 1.1 (no support to either of the model), 1.1 to 3.0 (positive support), 3.0 to 5.0 (strong support), and >5.0 (overwhelming support) were used to determine the best fit model. Coloured points represent provinces of sample collection, with point size scaled by the number of genomes contributing to the inference. Map Source: Statistics Canada Cartographic Boundary Files (https://www12.statcan.gc.ca/census-recensement/2021/geo/sip-pis/boundary-limites/index2021-eng.cfm?year=21)

## Discussion

This study reveals that influenza A(H3N2) subclade K detected in Canada were antigenically distinct, yet did not exhibit clear post-vaccine expansion during the early 2025/26 season, emphasizing that antigenic transition and epidemiologic growth are related but not synonymous processes [10,11]. These findings suggest that population immunity, seasonal timing, and geographical connectivity can modulate whether antigenic variants translate into increased transmission and spread [12]. This is supported by a recent Canadian interim study showing no substantial vaccine mismatch by HA inhibition assays and reported age-related variation in VE [5].

The phylogenetic structure of subclade K from Canada supports a highly connected interprovincial transmission network, characterized by frequent mixing and limited evidence of long-lived, province-specific lineage persistence. This pattern suggests that early epidemic dynamics were shaped by repeated introductions and re-seeding between jurisdictions, consistent with Canada functioning as a connected influenza metapopulation. Similar dynamics have been described for past clades of seasonal A(H3N2), in which viral persistence is maintained through ongoing migration rather than sustained local circulation [13]. Canadian syndromic and laboratory surveillance studies similarly support a coupled national pattern of influenza spread [14,15]. Within this context, phylogeographic reconstruction identified Ontario as a major source of interprovincial dissemination, likely reflecting its population size and connectivity, and highlighting the value of genomic surveillance in highly connected provinces for early situational awareness [16].

Sequence-based antigenic analyses indicate that subclade K occupies a distinct region of HA sequence space relative to J-lineage viruses and contemporary vaccine strains, supporting progressive divergence over successive seasons. Epitope-level analyses suggests that this divergence is concentrated at antigenically relevant sites (A and B), with elevated distances relative to prior seasons (NH 2022/23, NH 2023/24, SH 2023 and SH 2024) vaccine strains, consistent with reduced recognition by pre-existing immunity (Figure 2**B**, Supplementary Figures S2–S3). However, divergence relative to the NH 2025/26 and SH 2025 vaccine strains was moderate, supporting partial mismatch [17] rather than complete immune escape. The substantially lower antigenic distance observed relative to the SH 2026 vaccine strain suggests improved antigenic alignment and potentially greater preservation of vaccine-mediated protection depending on vaccine composition. The concentration of substitutions at or adjacent to predicted N-linked glycosylation motifs supports a plausible immune-evasion mechanism whereby AA replacement and altered glycan shielding jointly reduce antibody recognition [1,2,18,19].

These findings are subject to several limitations. Phylodynamic inference depends on the timing, density, and representativeness of genomic sampling; in Ontario, declining sequence availability toward December 2025 limited power to detect later-season transmission changes following the provincial activity peak, in late November to early December 2025. The R_t_ estimates reflect transmission among sampled lineages and may not capture heterogeneity across age groups, care settings, or communities. In addition, vaccine-associated effects cannot be disentangled from concurrent influences such as natural immunity, healthcare-seeking behavior, seasonal contact patterns, or vaccine uptake. Finally, uneven genomic surveillance across jurisdictions may obscure interprovincial linkages, highlighting the need for geographically balanced surveillance to assess delayed epidemiologic effects and ongoing viral evolution.

## Conclusion

Routine genomic surveillance integrated with antigenic assessment and phylogenetic modelling enables inference of transmission dynamics and geographic dissemination directly from pathogen sequence data, providing timely situational awareness to inform vaccine strain evaluation and public health decision-making.

## Data availability

All influenza A(H3N2) hemagglutinin (HA) sequences used in this study were retrieved from the GISAID database. The corresponding GISAID accession numbers are listed in Supplementary Excel file. Scripts are available at https://github.com/Phylodynamics-and-AI-for-Public-health/2025-2026EarlyKClade

## Authors’ contributions

Conceptualization: George S. Long, Samira Mubareka, Venkata R. Duvvuri; Data curation: George S. Long, Thomas W.A. Braukmann, Venkata R. Duvvuri; Formal Analysis and Interpretation: George S. Long, Thomas W.A. Braukmann, Samira Mubareka, Venkata R. Duvvuri; Methodology: George S. Long, Thomas W.A. Braukmann, Nicholas Waglechner, Patryk Aftanas, Shawn T. Clark, Finlay Maguire, Venkata R. Duvvuri; Visualization: George S. Long, Thomas W.A. Braukmann, Venkata R. Duvvuri; Sample collection coordination: Nicholas Waglechner, Patryk Aftanas, Alex Marchand-Austin; Supervision: Samira Mubareka, Venkata R. Duvvuri; Writing – original draft: George S. Long, Thomas W.A. Braukmann, Venkata R. Duvvuri; Writing – review & editing: George S. Long, Thomas W.A. Braukmann, Nicholas Waglechner, Patryk Aftanas, Alex Marchand-Austin, Julianne V. Kus, Shawn T. Clark, Kevin Katz, Maan Hasso, Finlay Maguire, Samir N. Patel, Samira Mubareka, Venkata R. Duvvuri

## Ethical statement

Ethical review was not needed for this study as it used de-identified publicly available data.

## Acknowledgements

We would like to thank the Biocomputing Center, Laboratory Surveillance and Data Management, and the Genomics Laboratory teams at Public Health Ontario for generating influenza sequence data used in this study. Sequences from Public Health Ontario were obtained as part of routine surveillance of human influenza viruses. Additional sequences were generated and shared in the public domain by the Shared Hospital Laboratory. We express our gratitude to Ruimin Gao and Nathalie Bastien of the Public Health Agency of Canada –National Microbiology Laboratory and researchers from the Saskatchewan Health Authority, the Roy Romanow Provincial Laboratory, and Hôpital Georges L. Dumont for generating and posting influenza sequences to the GISAID EpiFlu™ Database on which this research is based.

