## Supplementary Materials for "Genomic, antigenic and transmission dynamics of influenza A(H3N2) subclade K in Canada, early 2025/26 season"

### Supplementary Methods

#### Sequences and data sources

A total of 608 hemagglutinin (HA) and neuraminidase (NA) nucleotide sequences from influenza A(H3N2) clinical samples collected in Canada were retrieved from the GISAID database [1]. Sequences submitted between May 01, 2025, and January 06, 2026, were included to capture the inter-seasonal period and the early phase of the 2025/26 influenza season. Northern and Southern Hemisphere influenza A(H3N2) vaccine strains from 2022–2026 [2] were also included in the dataset.

Ontario sequences were generated by the Shared Hospital Laboratory (SHL) and Public Health Ontario (PHO). In total, 64 A(H3N2) sequences from PHO and 84 from SHL were available on GISAID. At PHO, samples were selected for sequencing based on the following criteria: (i) cycle threshold (Ct) value ≤ 27; (ii) nasopharyngeal or throat swabs with volumes ≥ 1 mL; (iii) absence of co-infections; and (iv) for outbreak-associated samples, sequencing of only the earliest representative specimen meeting these criteria., to avoid duplicate sampling from the same patient. At SHL, sequencing criteria were less restrictive; samples generally required (i) Ct ≤ 25, although some higher-Ct samples were included, and (ii) nasopharyngeal or throat swabs with volumes ≥ 0.5 mL. Following quality filtering (see below), 61 A(H3N2) sequences from PHO and 35 from SHL were retained for downstream analyses. Metadata for all samples included in this study are provided in the Supplementary spreadsheet.

#### Genome sequencing

**At PHO,** flu genomic segment sequences were generated by extracting total RNA with the MGISP-960 system using a MGI 960 extraction kit (MGI Tech, China). cDNA fragments were synthesized using the Superscript III one step RT PCR Kit with Platinum Taq polymerase (Thermo Fisher Scientific, USA) using the Opti1 primers (Opti1-F1 5’ GTTACGCGCCAGCAAAAGCAGG, Opti1-F2 5’GTTACGCGCCAGCGAAAGCAGG, Opti1-R1 5’GTTACGCGCCAGTAGAAACAAGG) [3]. RT-PCR cycling conditions used to generate fragments were one cycle of 55°C for 2 min, 42°C for 60 min, and 94°C for 2 min; followed by 5 cycles of 94°C for 30 sec, 44°C for 30 sec, and 68°C for 3 min 30 sec; followed by 35 cycles of 94°C for 30 sec, 57°C for 30 sec, 68°C for 3 min 30 sec; followed by a final extension of 68°C for 10 min. Prior to Illumina library preparation, samples were cleaned with AMPure XP beads (Beckman Coulter, USA) using a 1:0.9 bead to sample volume ratio. DNA concentration (ng/ul) was determined using the Qubit dsDNA High Sensitivity kit (Thermo Fisher Scientific, USA) on a Varioskan LUX Multimode Microplate Reader (Thermo Fisher Scientific, USA). Libraries for Illumina sequencing were generated using the Nextera XT DNA library prep kit (Illumina USA) following manufacturer's instructions for tagmentation and using Nextera DNA indices. Following library preparation, samples were cleaned using the same procedure as above, however, using a 1:0.8 bead to sample volume ratio. Samples were pooled in equimolar proportions and the final library fragment size checked on a 4200 TapeStation using a D1000 ScreenTape. Samples were sequenced on an Illumina NextSeq 550 system (Illumina, USA) using a NextSeq 550 Mid Output Kit V2 (Illumina, USA) with 302 cycles in paired-end mode (2×150 bp) with a 2% PhiX Control v2 (Illumina, USA) spiked into the final libraries.

**At SHL**, total RNA was extracted on a Hamilton Star (Hamilton Company, Reno, USA) automated liquid handling system using the Maxwell HT Viral TNA Kit (Promega, USA). Amplification was then performed using the SuperScript III One-Step RT-PCR System with Platinum Taq DNA Polymerase (Thermo Fisher Scientific, USA) with universal primers (Uni12/Inf-1 GGGGGGAGCAAAAGCAGG, Uni12/Inf-3 GGGGGGAGCGAAAGCAGG, and Uni13/Inf-1 CGGGTTATTAGTAGAAACAAGG) [4]. The RT-PCR thermal cycling profile consisted of reverse transcription at 42°C for 60 min and initial denaturation at 94°C for 2 min, followed by 5 cycles of 94°C for 30 s 44°C for 30 s, and 68°C for 3 min; followed by 35 cycles of 94°C for 30 s, 57°C for 30 s, and 68°C for 3 min; followed by a final extension at 68°C for 5 min. Following amplification, the resulting products were purified via automated SPRI bead cleanup (Illumina, USA) at a 1× ratio using an MGISP-960 platform (MGI Tech, China). DNA concentrations were quantified with the Quant-iT 1X dsDNA High Sensitivity Assay Kit (Thermo Fisher Scientific, USA) on a BioTek Synergy LX Multimode Reader (Agilent Technologies, USA). Sequencing libraries were generated using the DNA Prep Kit (Illumina, USA) with Unique Dual Indexes (Illumina) according to the manufacturer's protocol. Final libraries were quantified using the Quant-iT Assay, and pooled library size distribution was assessed on a 4150 TapeStation using a D1000 ScreenTape (Agilent Technologies, USA). Samples were normalized to equimolar concentrations and pooled for sequencing. The library pool was spiked with 2% PhiX Control v3 (Illumina, USA) and sequenced on an Illumina MiniSeq system (Illumina, USA) using a 300-cycle reagent kit in paired-end mode (2×149 bp).

Following sequencing, raw sequences are filtered using fastp v0.23.2 [5]. For both SHL and PHO, sequences were analysed using the nf-flu pipeline (v3.10.1) to generate consensus sequences, subtype prediction, and clade assignment using Nextclade [6,7]. All sequences are available on GISAID (Supplementary spreadheet)

### Sequence Quality Control and Alignment

The signal peptides were trimmed from the ends of the proteins and were subsequently deduplicated using the sequence content, date of collection, and geographic location. These sequences were then aligned using MAFFT v7.505 [8] with the results being further filtered to remove any strains consisting of ≥ 10% Ns or three consecutive gaps in their alignment. Following this, the intersection of the remaining HA, NA, and PA sequences were used in the remaining analyses to ensure that the same viral strains were included in both analyses. This led to a total of 480 Canadian H3N2 strains being included in this study. Vaccine strains used from the 2022/2023 to the upcoming 2026 flu seasons were also included for a total of 488 genomes. Sample metadata is available in the Supplementary spreadsheet.

**Phylogenetic, Phylodynamic and Phylogeographic Analysis**

A time-scaled phylogeny was created for all Canadian H3N2 sequences with the 2025-2026 Vaccine reference strains for the Southern Hemisphere. Sequences were aligned in Mafft v7.525 [8] using default parameters. A maximum likelihood tree then was inferred using IQ-TREE3 [9] using ModelFinder [10], 10 000 ultrafast bootstraps [11], and 10 000 aLRT calculations. Treetime v0.11.4 [12] was used to create a time-scaled phylogeny with default parametes using a clock rate inferred from TempEst and with stochastic resolve enabled. The resulting timestamped phylogeny was visualised in RStudio using R v4.5.1 with ape, ggtree, and treeio packages [13–17] (**Figure 1**).

The phylodynamics analysis was limited to only circulating genomes which are part of the K clade to ensure the tractability of the results. Maximum-likelihood trees were created using the same method as above, however, TempEST was used to identify outliers in the root-to-tip and residual plots (see the Supplementary spreadsheet for specific genomes) [18]. After filtering for outliers, the phylogenies were recreated and the clock rate estimated with TempEST.

Time-dependent reproductive numbers ($R_t$) were modelled using the Serial Birth-Death Skyline model implemented by BDSKY in Beast 2 [19,20] for the Canadian and Ontario subset of genomes. Twelve epochs were used to model the trajectory of $R_t$ in the Canadian model while the only 8 were used for the Ontario data set. The $R_t$ values were modelled using a log-normal distribution (*lognormal(1,1.25)*) bounded between 0 and 5. The uninfectiousness rate was fixed to 52, representing an infectious period of approximately one week, to ensure that the model would converge. The prior for the sampling rate was a log-normal distribution (*lognormal(0.1,1.25)*) bounded between 0 and 1. A strict clock rate was used based on the results found in the TempEst analysis and modeled using a log-normal distribution bounded between 0 and 0.01. The means of the distributions were determined based on the TempEst results. The BDSKY models were run using an adaptive Metropolis-Coupled Markov Chain Monte Carlo implemented in the CoupledMCMC package [21] with four chains for 60 million iterations over three runs.

A discrete trait diffusion phylogeographic analysis was performed with the Canadian dataset using BeastX [22]. Asymmetric transmission rates between the provinces were assumed and the strength of these transmission rates calculated using the Bayesian Stochastic Search Variable Selection (BSSVS) method. A constant population size model was used and with the same strict clock prior as in the BDSKY analysis. The model was run three times for 60 million iterations each. Once converged, the provincial rates were combined after using a 10% burn-in and the strengths estimated with SpreadD3 [23]. Transmission rates were interpreted using

$log_{10}(Bayes\ Factor)$: 0 to 1.1 (no support to either of the model), 1.1 to 3.0 (positive support), 3.0 to 5.0 (strong support), and >5.0 (overwhelming support) were used [24].

**Sequence- and Antigenic- Divergence Analysis**

Amino acid (AA) alignments of the HA and NA proteins were created with MAFFT [8] using the same strains as in the phylodynamic analysis. Segments from the relevant vaccine strains were also included in the alignment. Antigenic sites for H3 were identified by compiling the results of previous papers. The antigenic distances of the H3 segments were calculated using the FLU substitution matrix, which is based on the long-term evolution of Influenza [25], and used to perform an antigenic Principal Coordinates Analysis. Distances between the H3N2 vaccines were estimated with Bayesian regression modelling using the brms R package [26]. A lognormal prior was used to model the AA distances with the formula $Distance + 10^{-6} \sim Vaccine \cdot Clade$. The estimated marginal means of the model were then calculated ($\sim Vaccine \mid Clade$).

Alongside the FLU substitution matrix distances, the pEpitope distances to the vaccine strains were also calculated [27]. This was done by determining the number of mismatches between the antigenic sites (i.e. the Hamming distance) and dividing it by the total number of residues in the site. The global distance was then identified by selecting the most distant antigenic site. The same modelling structure used in the FLU substitution matrix was used here as well; however, a beta prior was used instead. Additionally, an antigen specific Bayesian regression model was run using the structure $Distance + 10^{-6} \sim Vaccine \cdot Clade \cdot Antigen$. The estimated marginal means of this model were calculated as $\sim Vaccine \mid Clade\ \cdot Antigen$. Glycosylation sites were identified using data from NextStrain [28].

### Supplementary Tables

Supplementary Table S1. **Estimated pEpitope distances between the vaccines and the H3 clades**. I: 22/23 NH; 23SH; 23/24 NH vaccine strains; II: 24 SH; 24/25 NH vaccine strains; III: 25 SH; 25/26 NH vaccine strains and IV: 26 SH vaccine strains

| H3 Clade | Vaccine | Median (95% HPD) |
|---|---|---|
| **J.2** | I | 0.149 [0.137, 0.161] |
| | II | 0.104 [0.094, 0.115] |
| | III | 0.084 [0.075, 0.094] |
| | IV | 0.008 [0.005, 0.010] |
| **J.2.1** | I | 0.167 [0.129, 0.206] |
| | II | 0.112 [0.081, 0.147] |
| | III | 0.112 [0.081, 0.146] |
| | IV | 0.112 [0.080, 0.144] |
| **J.2.2** | I | 0.176 [0.165, 0.187] |
| | II | 0.120 [0.111, 0.130] |
| | III | 0.102 [0.094, 0.111] |
| | IV | 0.121 [0.112, 0.130] |
| **J.2.3** | I | 0.163 [0.148, 0.179] |
| | II | 0.107 [0.094, 0.119] |
| | III | 0.078 [0.067, 0.090] |
| | IV | 0.109 [0.096, 0.122] |
| **J.2.4** | I | 0.190 [0.183, 0.196] |
| | II | 0.135 [0.129, 0.141] |
| | III | 0.135 [0.130, 0.141] |
| | IV | 0.002 [0.001, 0.002] |

| | | |
|---|---|---|
| **J.2.5** | I | 0.168 [0.152, 0.184] |
| | II | 0.113 [0.100, 0.126] |
| | III | 0.056 [0.046, 0.065] |
| | IV | 0.114 [0.101, 0.127] |
| **K** | I | 0.225 [0.223, 0.228] |
| | II | 0.170 [0.168, 0.172] |
| | III | 0.170 [0.168, 0.172] |
| | IV | 0.078 [0.077, 0.080] |

Supplementary Table S2. **Amino Acid substitutions identified in antigenic sites of the H3 hemagglutinin segment.** Only sites where the minor allele was present in more than one sample or is not solely present in a vaccine strain are included.

| Antigen | Position | AA | Vaccine | | | | 2a.3a.1 | | | | | | |
|---|---|---|---|---|---|---|---|---|---|---|---|---|---|
| | | | I | II | III | IV | J.2.1 | J.2 | J.2.2 | J.2.3 | J.2.4 | J.2.5 | K |
| A | 124 | N | - | - | - | - | - | 1 | 13 | - | - | - | 2 |
| | | S | 2 | 2 | 2 | 2 | 1 | 8 | - | 6 | 38 | 6 | 404 |
| | | X | - | - | - | - | - | - | - | - | 1 | - | - |
| | 135 | A | - | - | - | - | - | 1 | - | - | - | - | - |
| | | E | - | - | - | - | - | - | - | - | - | - | 1 |
| | | K | - | - | - | 2 | - | 1 | - | - | 33 | - | 405 |
| | | N | - | - | - | - | - | - | - | - | 6 | - | - |
| | | T | 2 | 2 | 2 | - | 1 | 7 | 13 | 6 | - | 6 | - |
| | 144 | D | - | - | - | - | - | - | - | - | - | - | 11 |
| | | N | - | - | - | - | - | - | - | - | 14 | - | 395 |
| | | S | 2 | 2 | 2 | 2 | 1 | 9 | 13 | 6 | 25 | 6 | - |
| | 145 | G | - | - | - | - | - | 1 | - | - | - | - | - |
| | | N | - | - | 2 | - | - | 2 | 2 | 5 | - | 6 | 1 |
| | | S | 2 | 2 | - | 2 | 1 | 6 | 11 | 1 | 39 | - | 405 |
| | 260 | I | 2 | 2 | 2 | 2 | 1 | 9 | 13 | 6 | 39 | 6 | 403 |
| | | V | - | - | - | - | - | - | - | - | - | - | 3 |
| | 261 | Q | - | - | - | - | 1 | 1 | - | - | - | - | 3 |
| | | R | 2 | 2 | 2 | 2 | - | 8 | 13 | 6 | 39 | 6 | 403 |
| B | 158 | D | - | - | - | - | - | - | - | - | 13 | - | 406 |
| | | K | - | - | - | - | - | - | - | 6 | - | 6 | - |
| | | N | 2 | 2 | 2 | 2 | 1 | 9 | 13 | - | 26 | - | - |
| | 159 | N | 2 | 2 | 2 | 2 | 1 | 9 | 12 | 6 | 39 | 6 | 404 |

| | | | | | | | | | | | | | |
|---|---|---|---|---|---|---|---|---|---|---|---|---|---|
| | | S | - | - | - | - | - | - | 1 | - | - | - | - |
| | | Y | - | - | - | - | - | - | - | - | - | - | 2 |
| | 160 | I | 2 | 2 | 2 | 2 | 1 | 9 | 13 | 6 | 26 | 6 | - |
| | | K | - | - | - | - | - | - | - | - | 13 | - | 405 |
| | | X | - | - | - | - | - | - | - | - | - | - | 1 |
| | 165 | D | - | - | - | - | - | - | - | - | - | - | 2 |
| | | K | - | - | - | - | - | - | - | - | 1 | - | 1 |
| | | N | 2 | 2 | 2 | 2 | 1 | 9 | 13 | 5 | 37 | 6 | 401 |
| | | X | - | - | - | - | - | - | - | 1 | 1 | - | 2 |
| | 189 | K | 2 | 2 | 2 | - | 1 | 7 | 13 | - | - | 6 | - |
| | | R | - | - | - | 2 | - | 2 | - | 6 | 39 | - | 406 |
| **C** | 46 | S | 2 | 2 | 2 | 2 | 1 | 9 | 13 | 6 | 39 | 6 | 404 |
| | | T | - | - | - | - | - | - | - | - | - | - | 1 |
| | | X | - | - | - | - | - | - | - | - | - | - | 1 |
| | 48 | I | 2 | 2 | 2 | 2 | 1 | 9 | 13 | 6 | 39 | 6 | 403 |
| | | T | - | - | - | - | - | - | - | - | - | - | 2 |
| | | X | - | - | - | - | - | - | - | - | - | - | 1 |
| | 54 | N | - | - | - | - | - | - | - | 5 | - | - | - |
| | | S | 2 | 2 | 2 | 2 | 1 | 9 | 13 | 1 | 39 | 6 | 405 |
| | | X | - | - | - | - | - | - | - | - | - | - | 1 |
| | 276 | E | - | - | 2 | 2 | 1 | 9 | 13 | 6 | 39 | 6 | 401 |
| | | K | 2 | 2 | - | - | - | - | - | - | - | - | 4 |
| | | V | - | - | - | - | - | - | - | - | - | - | 1 |
| | 278 | E | - | - | - | - | - | - | 1 | - | - | - | 12 |
| | | K | 2 | 2 | 2 | 2 | 1 | 9 | 12 | 6 | 39 | 6 | 394 |
| | 279 | F | - | - | - | - | - | - | - | - | 2 | - | - |
| | | S | 2 | 2 | 2 | 2 | 1 | 9 | 13 | 6 | 37 | 6 | 406 |

| | | | | | | | | | | | | | |
|---|---|---|---|---|---|---|---|---|---|---|---|---|---|
| | 309 | I | - | - | - | - | - | - | - | 1 | - | - | 2 |
| | | V | 2 | 2 | 2 | 2 | 1 | 9 | 13 | 5 | 39 | 6 | 404 |
| | 96 | I | - | - | - | - | - | 1 | - | - | - | - | - |
| | | N | 2 | - | - | - | - | - | - | - | - | - | 1 |
| | | S | - | 2 | 2 | 2 | 1 | 8 | 13 | 6 | 39 | 6 | 405 |
| | 173 | Q | 2 | 2 | 2 | 2 | 1 | 9 | 13 | 6 | 39 | 6 | 1 |
| | | R | - | - | - | - | - | - | - | - | - | - | 405 |
| | 214 | I | 2 | 2 | 2 | 2 | - | 9 | 10 | 6 | 38 | 6 | 405 |
| | | S | - | - | - | - | - | - | - | - | - | - | 1 |
| | | T | - | - | - | - | 1 | - | 3 | - | 1 | - | - |
| | 216 | H | - | - | - | - | - | - | - | 2 | - | - | - |
| | | N | 2 | 2 | 2 | 2 | - | 9 | 13 | 4 | 39 | 6 | 406 |
| | | S | - | - | - | - | 1 | - | - | - | - | - | - |
| | 221 | P | 2 | 2 | 2 | 2 | 1 | 9 | 13 | 6 | 39 | 6 | 401 |
| **D** | | X | - | - | - | - | - | - | - | - | - | - | 5 |
| | 222 | K | - | - | - | - | - | - | - | - | - | - | 2 |
| | | R | 2 | 2 | 2 | 2 | 1 | 8 | 13 | 6 | 38 | 6 | 401 |
| | | X | - | - | - | - | - | 1 | - | - | 1 | - | 3 |
| | 229 | G | - | - | - | - | - | - | - | - | - | - | 1 |
| | | I | - | - | - | - | - | - | 1 | - | - | - | - |
| | | K | - | - | - | - | - | - | - | - | - | - | 1 |
| | | R | 2 | 2 | 2 | 2 | 1 | 9 | 12 | 6 | 39 | 6 | 402 |
| | | X | - | - | - | - | - | - | - | - | - | - | 2 |
| | 230 | I | 2 | 2 | 2 | 2 | 1 | 9 | 13 | 6 | 39 | 6 | 404 |
| | | V | - | - | - | - | - | - | - | - | - | - | 2 |
| | 242 | I | 2 | 2 | 2 | 2 | 1 | 9 | 12 | 6 | 38 | 6 | 404 |
| | | M | - | - | - | - | - | - | - | - | 1 | - | 2 |

| | | | | | | | | | | | | | |
|---|---|---|---|---|---|---|---|---|---|---|---|---|---|
| | | T | - | - | - | - | - | - | 1 | - | - | - | - |
| | 244 | L | 2 | 2 | 2 | 2 | 1 | 9 | 11 | 6 | 39 | 6 | 406 |
| | | V | - | - | - | - | - | - | 2 | - | - | - | - |
| | 246 | I | - | - | - | - | - | - | - | - | - | - | 1 |
| | | K | - | - | - | - | - | - | - | - | - | - | 1 |
| | | N | 2 | 2 | 2 | 2 | 1 | 9 | 12 | 6 | 37 | 6 | 403 |
| | | X | - | - | - | - | - | - | 1 | - | 2 | - | 1 |
| | 59 | I | - | - | - | - | - | - | - | - | - | - | 2 |
| | | L | 2 | 2 | 2 | 2 | 1 | 9 | 13 | 6 | 39 | 6 | 403 |
| | | X | - | - | - | - | - | - | - | - | - | - | 1 |
| | 62 | G | 2 | 2 | 2 | 2 | 1 | 9 | 11 | 6 | 39 | 6 | 405 |
| | | R | - | - | - | - | - | - | 2 | - | - | - | - |
| | | X | - | - | - | - | - | - | - | - | - | - | 1 |
| | 63 | D | - | - | - | - | - | - | - | 1 | - | - | - |
| | | N | 2 | 2 | 2 | 2 | 1 | 9 | 13 | 5 | 39 | 6 | 400 |
| | | S | - | - | - | - | - | - | - | - | - | - | 4 |
| **E** | | X | - | - | - | - | - | - | - | - | - | - | 2 |
| | 67 | I | 2 | 2 | 2 | 2 | 1 | 5 | 13 | 6 | 39 | 6 | 405 |
| | | M | - | - | - | - | - | 4 | - | - | - | - | - |
| | | X | - | - | - | - | - | - | - | - | - | - | 1 |
| | 78 | G | 2 | 2 | 2 | 2 | 1 | 5 | 13 | 4 | 39 | 6 | 406 |
| | | S | - | - | - | - | - | 4 | - | 2 | - | - | - |
| | 83 | D | - | - | - | - | - | - | 2 | - | - | - | - |
| | | E | 2 | 2 | 2 | 2 | 1 | 9 | 11 | 6 | 39 | 6 | 406 |
| | 260 | I | 2 | 2 | 2 | 2 | 1 | 9 | 13 | 6 | 39 | 6 | 403 |
| | | V | - | - | - | - | - | - | - | - | - | - | 3 |
| **RBS** | 135 | A | - | - | - | - | - | 1 | - | - | - | - | - |

| | | | | | | | | | | | | |
|---|---|---|---|---|---|---|---|---|---|---|---|---|
| | E | - | - | - | - | - | - | - | - | - | - | 1 |
| | K | - | - | - | 2 | - | 1 | - | - | 33 | - | 405 |
| | N | - | - | - | - | - | - | - | - | 6 | - | - |
| | T | 2 | 2 | 2 | - | 1 | 7 | 13 | 6 | - | 6 | - |
| 158 | D | - | - | - | - | - | - | - | - | 13 | - | 406 |
| | K | - | - | - | - | - | - | - | 6 | - | 6 | - |
| | N | 2 | 2 | 2 | 2 | 1 | 9 | 13 | - | 26 | - | - |
| 159 | N | 2 | 2 | 2 | 2 | 1 | 9 | 12 | 6 | 39 | 6 | 404 |
| | S | - | - | - | - | - | - | 1 | - | - | - | - |
| | Y | - | - | - | - | - | - | - | - | - | - | 2 |
| 221 | P | 2 | 2 | 2 | 2 | 1 | 9 | 13 | 6 | 39 | 6 | 401 |
| | X | - | - | - | - | - | - | - | - | - | - | 5 |
| 222 | K | - | - | - | - | - | - | - | - | - | - | 2 |
| | R | 2 | 2 | 2 | 2 | 1 | 8 | 13 | 6 | 38 | 6 | 401 |
| | X | - | - | - | - | - | 1 | - | - | 1 | - | 3 |
| 229 | G | - | - | - | - | - | - | - | - | - | - | 1 |
| | I | - | - | - | - | - | - | 1 | - | - | - | - |
| | K | - | - | - | - | - | - | - | - | - | - | 1 |
| | R | 2 | 2 | 2 | 2 | 1 | 9 | 12 | 6 | 39 | 6 | 402 |
| | X | - | - | - | - | - | - | - | - | - | - | 2 |
| 230 | I | 2 | 2 | 2 | 2 | 1 | 9 | 13 | 6 | 39 | 6 | 404 |
| | V | - | - | - | - | - | - | - | - | - | - | 2 |

Supplementary Table S3. **Glycosylation patterns of the H3 HA segment during the early 2025/2026 influenza season.** Sites in bold indicate gain of glycosylation sites related to the vaccine strains. The three letter codes indicate the specific AA residues of the N-glycosylation site.

| Clade | Position | | | | | | | | | | | | | Genomes |
|---|---|---|---|---|---|---|---|---|---|---|---|---|---|---|
| | 8 | 22 | 38 | 45 | 63 | 94 | 122 | 135 | 144 | 154 | 165 | 246 | 285 | |
| **I** | NST | NGT | NAT | NSS | NCT | - | NES | - | - | NET | NVT | NST | NGS | 2 |
| **II** | NST | NGT | NAT | NSS | NCT | NSS | NES | - | - | NET | NVT | NST | NGS | 2 |
| **III** | NST | NGT | NAT | NSS | NCT | NSS | - | - | - | NET | NVT | NST | NGS | 2 |
| **IV** | NST | NGT | NAT | NSS | NCT | NSS | - | - | - | NET | NVT | NST | NGS | 2 |
| **J.2** | - | NGT | NAT | NSS | NCT | NSS | - | - | - | NET | NVT | NST | NGS | 4 |
| | - | NGT | NAT | NSS | NCT | - | - | - | - | NET | NVT | NST | NGS | 2 |
| | NST | NGT | NAT | NSS | NCT | NSS | - | - | - | NET | NVT | NST | NGS | 2 |
| | NST | NGT | NAT | NSS | NCT | NSS | - | - | - | NET | NVT | NST | NGS | 1 |
| **J.2.1** | NST | NGT | NAT | NSS | NCT | NSS | - | - | - | NET | NVT | NST | NGS | 1 |
| | NST | NGT | NAT | NSS | - | NSS | - | - | - | NET | NVT | NST | NGS | 8 |
| | **NGT** | NGT | NAT | NSS | - | NSS | - | - | - | NET | NVT | NST | NGS | 2 |
| | NST | NGT | NAT | NSS | NCT | NSS | - | - | - | NET | NVT | NST | NGS | 2 |
| | NST | NGT | NAT | NSS | - | NSS | - | - | - | NET | NVT | - | NGS | 1 |
| **J.2.3** | NST | NGT | NAT | NSS | NCT | NSS | - | - | - | NET | NVT | NST | NGS | 4 |
| | - | NGT | NAT | NSS | - | NSS | - | - | - | NET | NVT | NST | NGS | 1 |
| | NST | NGT | NAT | NSS | NCT | NSS | - | - | - | NET | - | NST | NGS | 1 |
| **J.2.4** | NST | NGT | NAT | NSS | NCT | NSS | - | - | - | NET | NVT | NST | NGS | 19 |
| | NST | NGT | NAT | NSS | NCT | NSS | - | - | **NSS** | NET | NVT | NST | NGS | 14 |
| | NST | NGT | NAT | NSS | NCT | NSS | - | **NSS** | - | NET | NVT | NST | NGS | 2 |
| | NST | NGT | NAT | NSS | NCT | NSS | - | **NSS** | - | NET | - | NST | NGS | 2 |
| | NST | NGT | NAT | NSS | NCT | NSS | - | **NSS** | - | NET | - | - | NGS | 2 |
| **J.2.5** | NST | NGT | NAT | NSS | NCT | NSS | - | - | - | NET | NVT | NST | NGS | 6 |

| | | | | | | | | | | | | | | |
|---|---|---|---|---|---|---|---|---|---|---|---|---|---|---|
| | NST | NGT | NAT | NSS | NCT | NSS | - | - | **NSS** | NET | NVT | NST | NGS | 357 |
| | NST | NGT | NAT | NSS | NCT | NSS | - | - | **NSS** | NET | - | NST | NGS | 23 |
| | NST | NGT | NAT | NSS | NCT | NSS | - | - | **-** | NET | NVT | NST | NGS | 11 |
| | NST | NGT | NAT | NSS | - | NSS | - | - | **NSS** | NET | NVT | NST | NGS | 5 |
| | NST | NGT | NAT | NSS | NCT | NSS | - | - | **NSS** | NET | NVT | - | NGS | 2 |
| **K** | NST | NGT | NAT | NSS | NCT | NSS | - | - | **NSS** | NET | NVT | NST | NGS | 2 |
| | **NIT** | NGT | NAT | NSS | NCT | NSS | - | - | **NSS** | NET | NVT | NST | NGS | 1 |
| | NST | NGT | NAT | NSS | NCT | - | - | - | **NSS** | NET | NVT | NST | NGS | 1 |
| | NST | NGT | NAT | NSS | NCT | NSS | - | - | **NNS** | NET | NVT | NST | NGS | 1 |
| | NST | NGT | NAT | NSS | NCT | NSS | - | - | **NSS** | NET | - | - | NGS | 1 |
| | NST | NGT | NAT | NTS | NCT | NSS | - | - | **NSS** | NET | NVT | NST | NGS | 1 |
| | NST | NGT | - | - | - | NSS | - | - | **NSS** | NET | NVT | NST | NGS | 1 |

Supplementary Table S4. **Estimated time-varying reproduction numbers (Rt)for Ontario and Canada during the early 2025/2026 influenza season.**

| Province | Period | Median (95% HPD) |
|---|---|---|
| **Ontario** | August 18th – August 29th | 0.593 [0.024, 2.419] |
| | August 29th – September 9th | 1.556 [0.036, 3.798] |
| | September 9th – September 20th | 1.852 [0.113, 3.713] |
| | September 20th – October 1st | 1.394 [0.212, 2.684] |
| | October 1st – October 12th | 0.531 [0.046, 1.307] |
| | October 12th – October 23rd | 1.114 [0.250, 1.982] |
| | October 23rd – November 3rd | 2.767 [2.128, 3.410] |
| | November 3rd – November 14th | 0.255 [0.034, 0.594] |
| **Canada** | August 12th – August 23rd | 0.681 [0.026, 2.166] |
| | August 23rd – September 2nd | 0.971 [0.051, 2.562] |
| | September 2nd – September 13th | 1.160 [0.054, 2.685] |
| | September 13th – September 23rd | 1.087 [0.063, 2.390] |
| | September 23rd – October 4th | 1.148 [0.283, 2.137] |
| | October 4th – October 14th | 0.808 [0.104, 1.798] |
| | October 14th – October 25th | 2.079 [1.466, 2.726] |
| | October 25th – November 4th | 1.481 [0.998, 1.924] |
| | November 4th – November 15th | 0.769 [0.482, 1.083] |
| | November 15th – November 25th | 1.211 [0.900, 1.513] |
| | November 25th – December 6th | 0.403 [0.120, 0.720] |
| | December 6th – December 17th | 1.291 [0.794, 1.761] |

**Supplementary Figures**

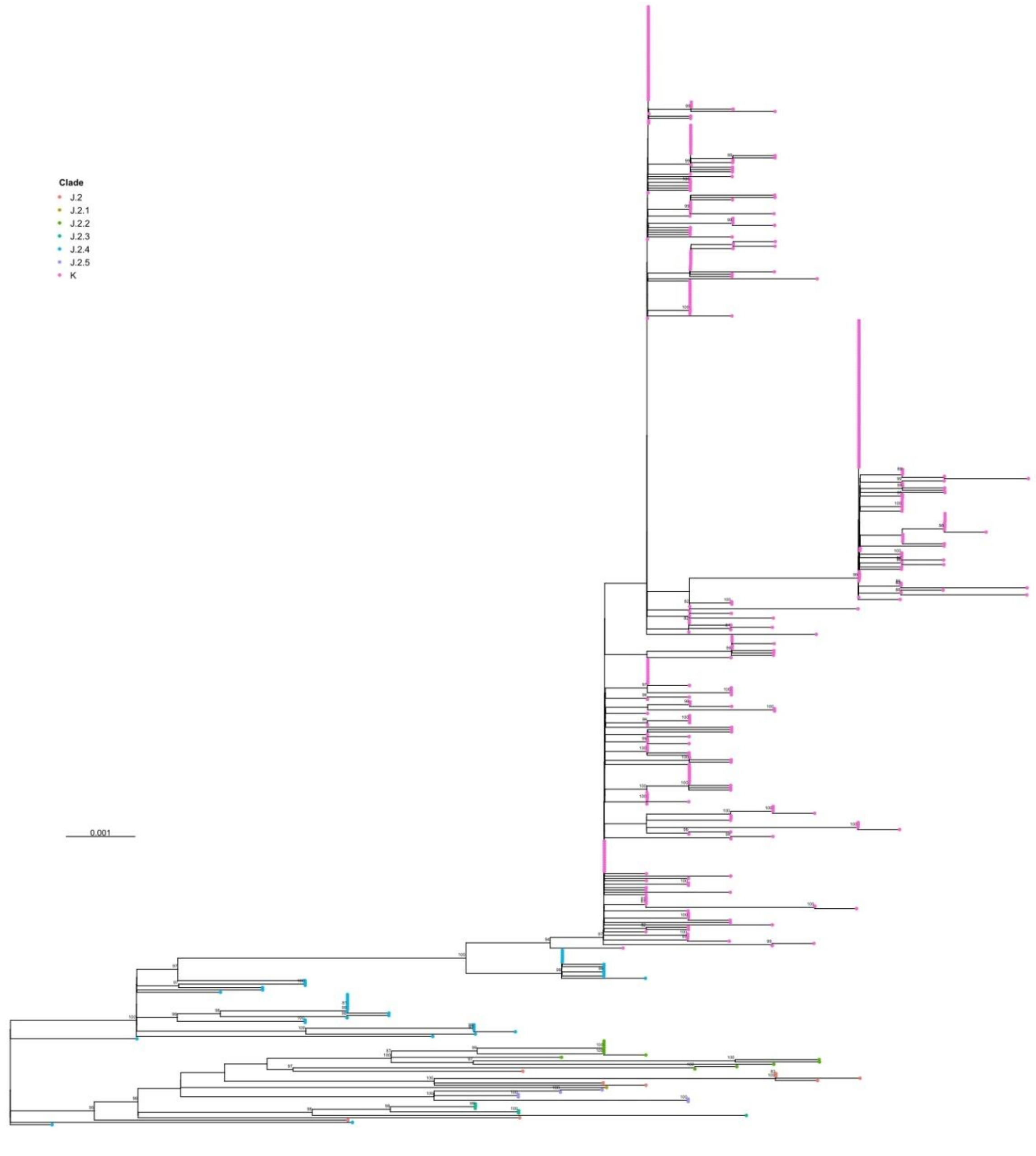

Supplementary Figure S1. **Time-scaled maximum-likelihood phylogeny of Canadian influenza A(H3N2) HA sequences (2025/26), including vaccine reference strains.** A time-aware maximum-likelihood phylogeny was inferred from HA alignments and subsequently time-scaled. Tips are coloured by clade (J.2, J.2.1, J.2.2, J.2.3, J.2.4, J.2.5, and K), and the x-axis represents time in years. Node support values (ultrafast bootstrap) are shown for internal branches where available. Vaccine reference strains are included for context.

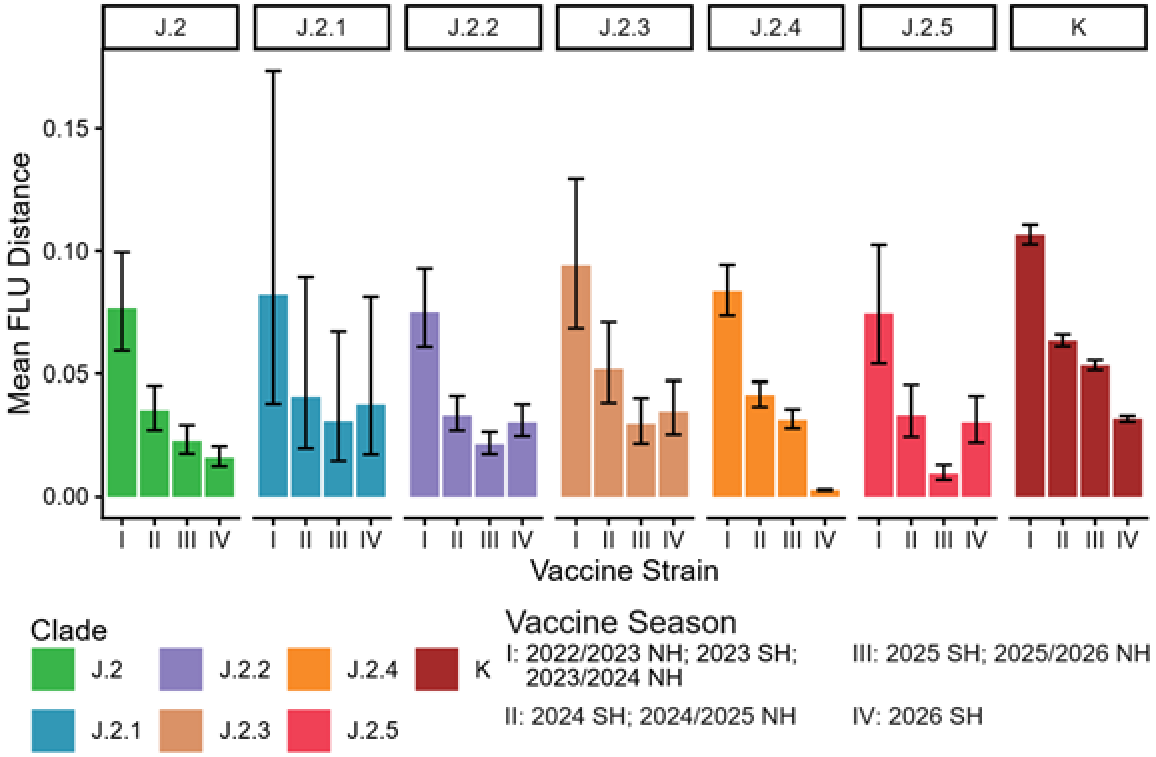

Supplmentary Figure S2. **HA Antigenic Site Divergence from Vaccine Strains Across Canadian A(H3N2) Subclades using the FLU Distance Model.** Mean FLU model distances between Canadian A(H3N2) HA sequences and vaccine reference strains, stratified by J-lineage subclade (J.2, J.2.1–J.2.5) and subclade K. Bars show mean distance and error bars indicate variability across sequences within each subclade. Vaccine strains are grouped by season: I (2022/23 NH; 2023 SH; 2023/24 NH), II (2024 SH; 2024/25 NH), III (2025 SH; 2025/26 NH), and IV (2026 SH).

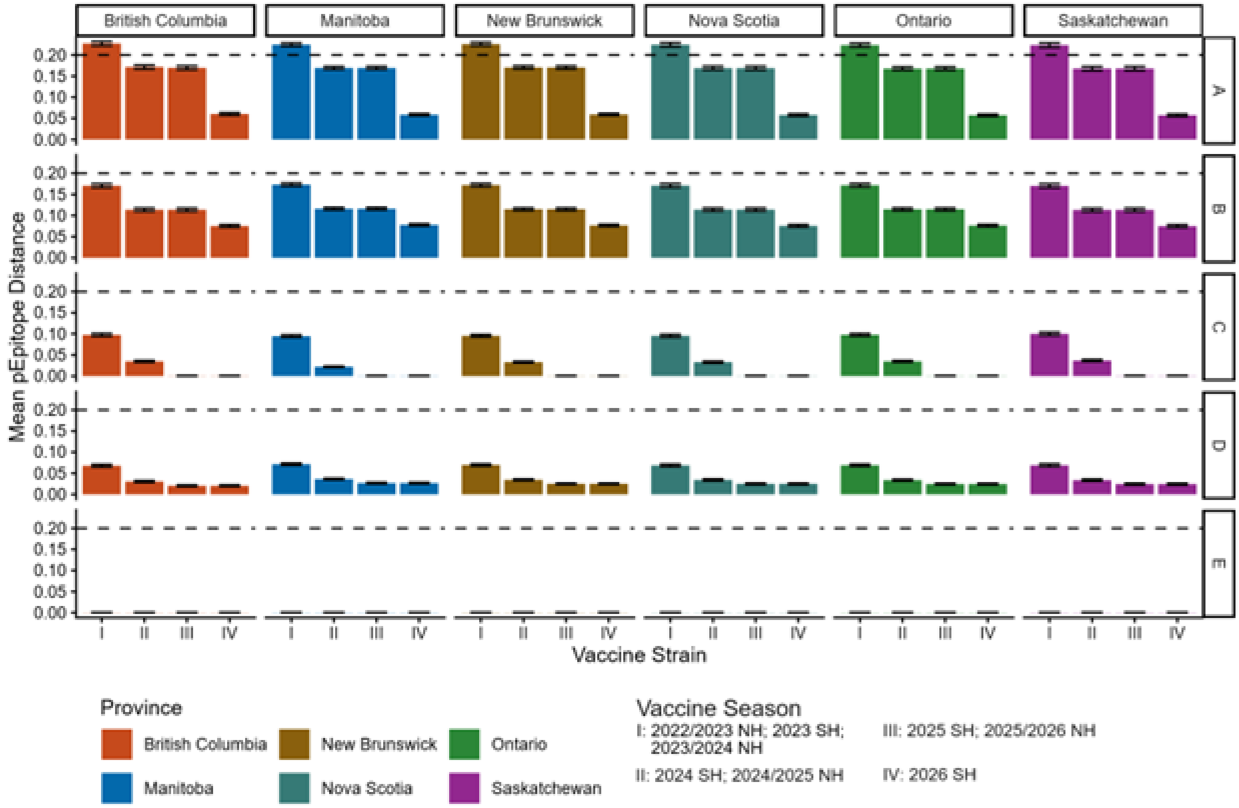

Supplementary Figure S3. **Province-specific pEpitope distances between Canadian A(H3N2) subclade K HA sequences and vaccine reference strains across five HA antigenic sites.** Mean pEpitope distances were calculated for subclade K sequences from British Columbia (BC), Manitoba (MB), New Brunswick (NB), Nova Scotia (NS), Ontario (ON), and Saskatchewan (SK), relative to four vaccine reference strains (I–IV). Panels A–E correspond to the five canonical HA antigenic sites (A, B, C, D, and E). Bars represent mean pEpitope with their 95% highest posterior densities for each province–vaccine comparison. The dashed horizontal line indicates the commonly used pEpitope threshold (pEpitope ≥ 0.20) [26] associated with substantial antigenic mismatch. Colours denote province.
